# Primary Care Physicians’ Practices and Barriers in Evaluating and Managing Chronic Kidney Disease in New Providence, The Bahamas

**DOI:** 10.1101/2024.09.13.24313661

**Authors:** W Bain, S Pinder-Butler, T Fountain, I Grant

## Abstract

**Objectives:** To assess the practices and barriers in evaluating and managing chronic kidney disease among primary care physicians in New Providence, The Bahamas.

**Methods:** A cross-sectional study utilizing an anonymous, self-administered questionnaire was given to General Practitioners, Family Medicine, and Internal Medicine physicians after using a simple random sampling approach. Descriptive and inferential statistical analysis was conducted using IBM SPSS software.

**Results:** There were 119 physicians in this study with Family Medicine specialty area representing 52.1%. Seventy-four (74) physicians reported following CKD guidelines. The most common at-risk groups identified were Diabetes Mellitus (100%), Hypertension (98.3%), and use of nephrotoxic agents (97.5%). The most common diagnostic test used to identify CKD was eGFR (97.5%) and 72.2% of physicians used eGFR alone to stage CKD. Physicians overall agreed (40.3 – 50.4%) they were comfortable in diagnosing and managing CKD and its complications except for bone disorders (43.2%) and metabolic acidosis (34.7%) where responses were neutral. Physicians were neutral in having tools/resources to help them manage bone disorders (35.3%) and metabolic acidosis (31.9%) and disagreed to having educational tools for patients to understand bone disorders (32.2%) and metabolic acidosis (32.8%). Physicians agreed-strongly agreed with 12 of 13 perceived barriers, and there were 26 unique barriers expressed (8 patient-level, 7 provider-level, 11 systems-level).

**Conclusions:** Deficits in the evaluation and management of CKD, and numerous barriers to CKD care were discovered. Recommendations include the development of a national CKD guideline, local CKD continuous medical education seminars, and public health campaigns on CKD education.

## Introduction

Chronic kidney disease (CKD) is a global public health concern. A systematic analysis of the Global Burden of Disease Study 2017 recorded 697.5 million cases of all-stage CKD, resulting in a global prevalence of ∼10%. In 2017, 1.2 million people died from CKD. There was an estimated 4.3 million cases of all-stage CKD with 11,023 deaths from 18 English and non-English speaking Caribbean countries in 2017. Of that, The Bahamas had an estimated 35,741 cases of all-stage CKD with 93 deaths that year.^1^ In the United States, the Centers for Disease Control and Prevention state that as many as 9 in 10 adults with CKD do not know they have it.^2^ CKD is expected to become the 5^th^ leading cause of mortality by 2040.^3^ Despite CKD being a global health concern and a risk factor for cardiovascular disease, primary care physicians (PCP) have raised several concerns which include, difficulties associated with assigning a diagnosis, the stigmatizing effect of a diagnosis, explaining the concept of CKD to patients, achieving blood pressure targets, complicated regimes, and uncertainty about referral to secondary care.^4^ This study aimed to assess the practices and barriers in evaluating and managing CKD among PCPs in New Providence, The Bahamas.

## Methods

This cross-sectional quantitative study was conducted on 119 out of 135 (response rate 88.1%) PCPs from October 2022 – February 2023 in New Providence, The Bahamas. Eligible PCPs included licensed General Practitioners, Family Medicine and Internal Medicine physicians working at community clinics and private clinics/offices. Physicians were excluded if they were unlicensed, working outside the areas of the study population, had sub-specialty training, predominantly caring for a specific population group (e.g., children, pregnant women), not working in New Providence, primarily hospital-based, and working in administrative roles. A simple random sampling approach was used on the list of 174 eligible physicians. The minimal sample size using Cochran’s formula, adjusting for the finite population of 174 physicians, and accounting for a 10% refusal was calculated to be 134. An anonymous, self-administered, 33-item questionnaire was used in this study and was created using modified questions from similar studies^5,6,10,11^. For face and content validity, the research team’s Epidemiologist and Nephrologist reviewed the questionnaire and agreed that the combined questions would measure the study objectives and meet target characteristics. Pre-testing was done from September – October 2022 on 10 physicians from the randomized sample frame and these physicians were not re-used in the study.

IBM SPSS Statistical Analysis application software (v 16) was used for descriptive and inferential statistical analysis. Descriptive statistics included frequencies, and the mean with standard deviation for ratio data (age in years and years in practice). Inferential statistics included the estimating of 95% confidence intervals around point estimates of interest in this study and included hypothesis testing of differences of interest assuming a Type 1 error of ± 5% (α is ± 0.05) using Pearson’s Chi-squared test. Based on smaller numbers of groups within medical training, variables were collapsed and used to determine statistical significance among the dependent variables of interest. For medical training physicians with a Bachelor of Medicine, Bachelor of Surgery (MBBS) and Medical Doctor (MD) degrees were combined, physicians with a Diploma and Master of Science in Family Medicine were combined, and physicians with Doctor of Medicine in Family Medicine and Internal Medicine were combined (MBBS/MD vs Diploma/MSc in FM vs DM in FM/IM). The qualitative data from the questionnaire was transferred to Microsoft Word to tally responses and to create themes for the unique barriers expressed.

This study was projected to pose minimal risk to the participants. Local International Review Board approval from the joint Public Hospitals Authority and University of the West Indies Ethics committee was granted on 18^th^ July 2022, and from the Ministry of Health Medical Research Oversight System on 16^th^ September 2022.

## Results

One hundred and thirty-five (135) physicians were sampled, and 119 questionnaires were completed (response rate = 88.1%). The demographic profile of the participating physicians is outlined in Table S1, Supplemental Material. The physicians were mostly female (63.9%, n = 76), Black/Afro-Caribbean (91.5%, n = 108), mean age 45.1 ± 10.1 years, and working within the Family Medicine specialty area (52.1%, n = 62). The most common highest level of completed medical training was the MBBS degree (31.4%, n = 37), followed by Doctor of Medicine (DM) in Family Medicine (28.8%, n = 34). For practice setting, “private practice only” was common (36.1%, n = 43), followed by physicians in both public and private practice (34.5%, n = 41) at approximately one-third. The mean years in practice was 15.8 ± 10.3 years. Regarding the number of patients seen per week, 51.3% (n = 61) of physicians reported seeing 51-100 patients per week, and 73.7% (n = 87) reported seeing 10 or less CKD patients per week. Of the 109 physicians who responded to whether they followed CKD guidelines, 67.9% (n = 74) reported that they did. Fifty-two (52) physicians specified 8 types of guidelines with KDIGO (Kidney Disease: Improving Global Outcomes) being the most frequently cited at 39.7% (n = 23). Based on medical specialty area, 48.9% (n = 22) of physicians in General Practice, 80.7% (n = 46) in Family Medicine, and 85.7% (n = 6) in Internal Medicine reported following CKD guidelines.

The 3 most common at-risk groups of developing CKD physicians identified were people with Diabetes Mellitus (100%, n = 118), Hypertension (98.3%, n = 116), and use of nephrotoxic agents (97.5%, n = 115), see Figure 1 below. The 3 least commonly selected at-risk groups were Elderly (66.9%, n = 79), Smoking (69.5%, n = 82), and Obesity (71.8%, n = 84). The most common diagnostic test physicians used to identify a patient with CKD was estimated glomerular filtration rate (eGFR) at 97.5%, n = 116, followed by ultrasound scan of the kidneys (73.1%, n = 87), isolated serum creatinine (65.5%, n = 78), 24-hour creatinine clearance with total protein (62.2%, n = 74), urinalysis (54.6%, n = 65), random urine microalbumin (52.1%, n = 62), random urine protein (37.8%, n = 45), and random urine creatinine (26.9%, n = 32), see Figure 2 below. Three-quarters of physicians (75.6 %, n = 90) reported requesting diagnostic tests in patients considered at risk. When CKD was not diagnosed in at-risk individuals during the first assessment, physicians reported repeating diagnostic tests annually (26.3%, n = 31), biannually (26.3%, n = 31), and at clinical discretion (19.5%, n = 23). Most physicians (82.2%, n = 97) classified the stage of CKD with eGFR alone (72.2%, n = 70) and 16.5% (n = 16) used eGFR and uACR (urine albumin-to-creatinine ratio). Approximately three-quarters of the physicians used 1 or more equations to calculate eGFR, with 40.3% (n = 48) reported use of the Chronic Kidney Disease Epidemiology Collaboration (CKD-EPI) equation, 32.8% (n = 39) the Modification of Diet in Renal Disease (MDRD) equation, and 17.6% (n = 21) relied on automated eGFR reporting by the lab.

**Figure 1:**
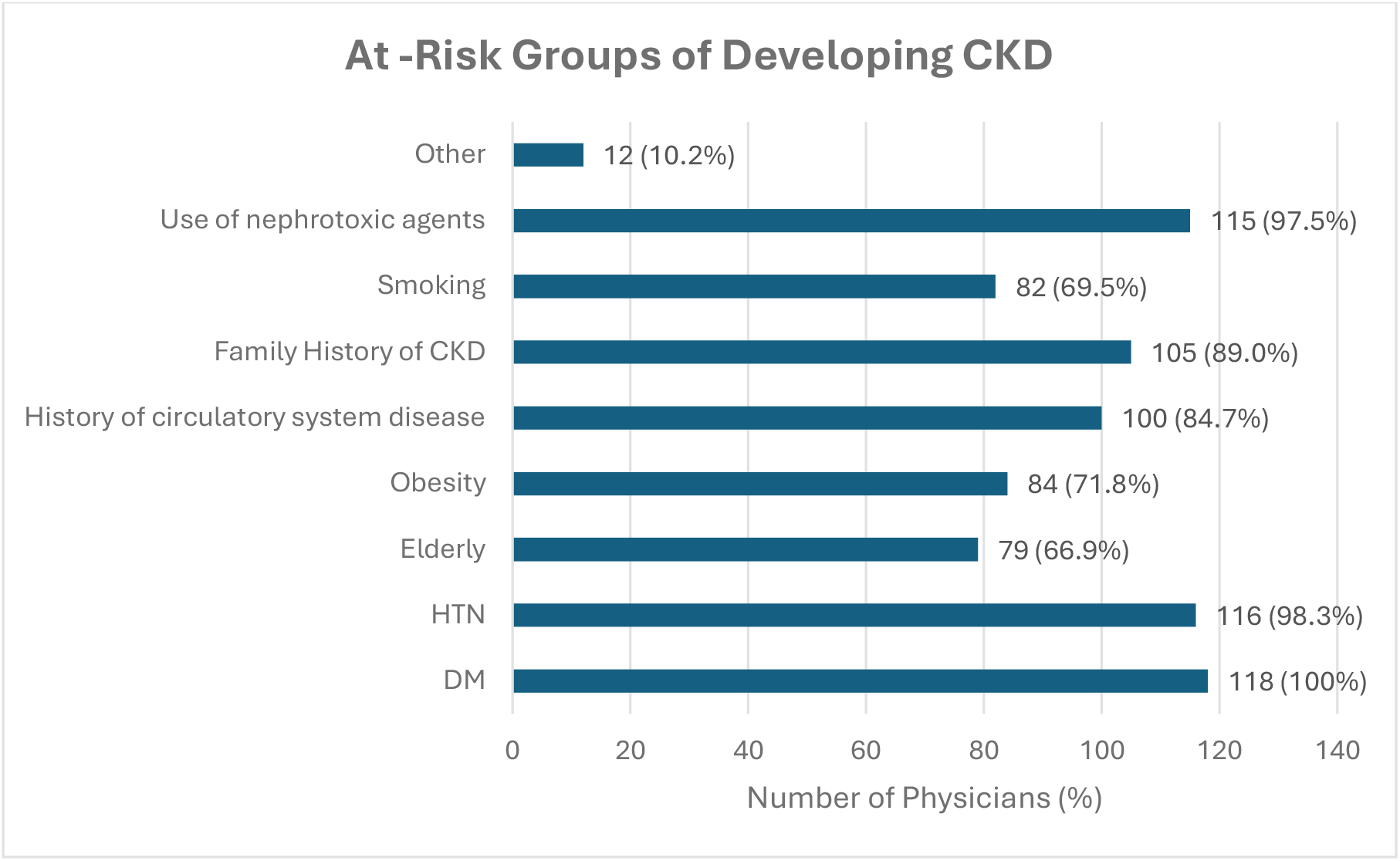
Physician Responses to Groups At-Risk of Developing CKD

**Figure 2:**
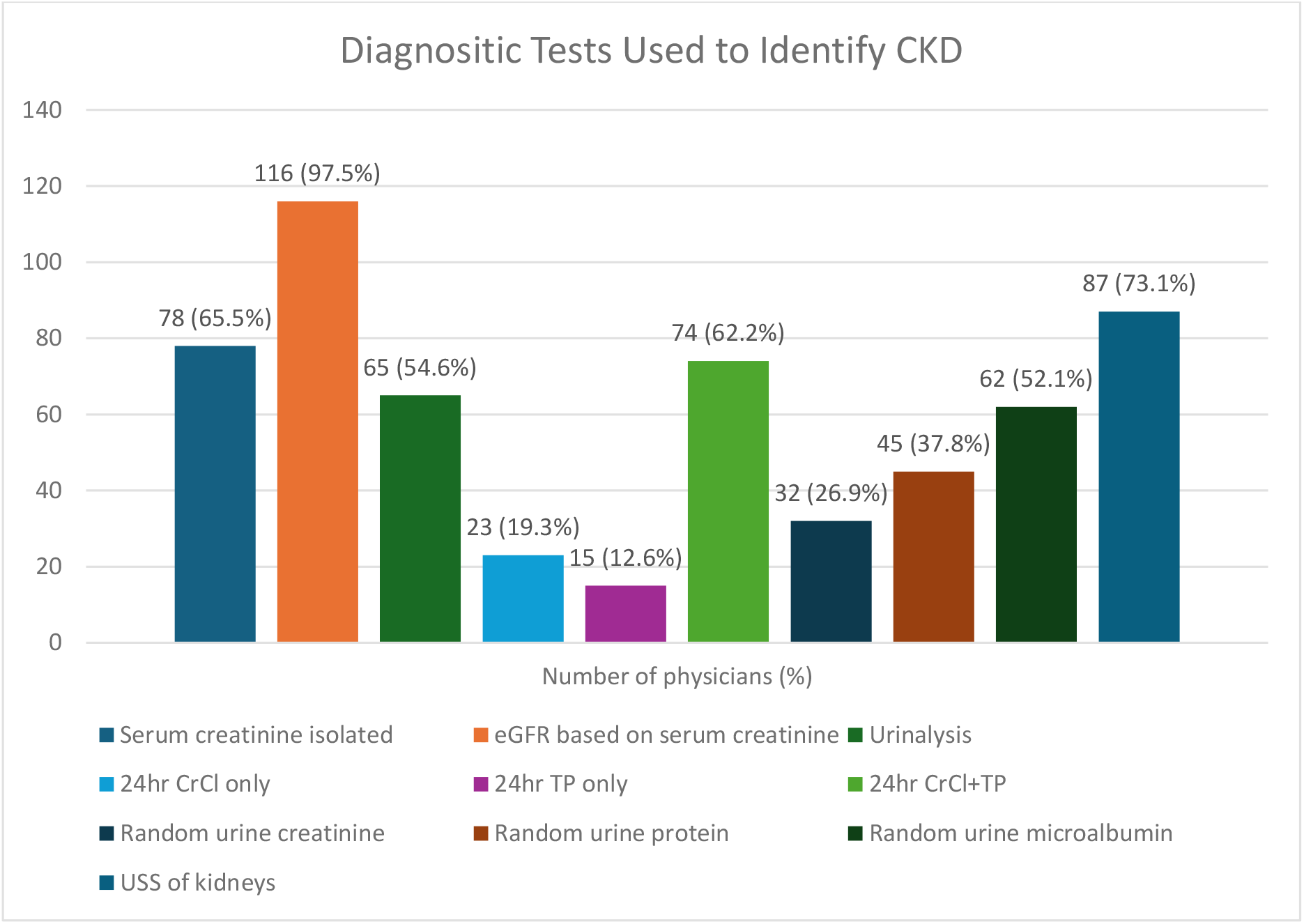
Diagnostic Tests Used by Physicians to Identify Patients with CKD Abbreviations: Chronic kidney disease (CKD); hypertension (HTN), diabetes mellitus (DM) estimated glomerular filtration rate (eGFR); creatinine clearance (CrCl); total protein (TP): ultrasound scan (USS)

Physicians were assessed using a 5-point Likert scale in comfort with various aspects of diagnosing and managing CKD, see Table S2.1 in Supplemental Material. Physicians “agreed” with feeling comfortable diagnosing, educating patients, managing medication dosing, avoiding nephrotoxic drugs, managing anemia and electrolyte disorders. For managing bone disorders and metabolic acidosis, responses were mostly “neutral” at 43.2% (n = 51) and 34.7% (n = 41), respectively. An assessment on the availability of tools revealed mostly “neutral” responses for tools to help physicians manage bone disorders (35.3%, n = 42) and metabolic acidosis (31.9%, n = 38) in CKD, and physicians “disagreed” in having tools to help their patients with bone disorders (32.2%, n = 38) and metabolic acidosis (32.8%, n = 39), see Tables S2.2-2.3 in Supplemental Material.

A 5-point Likert scale was used to assess confidence in monitoring and managing various aspects of CKD. For using urine protein results to manage CKD, physicians were equally “neutral” and “confident” at 33.1% (n = 39), see Table 1 below.

**Table 1:**
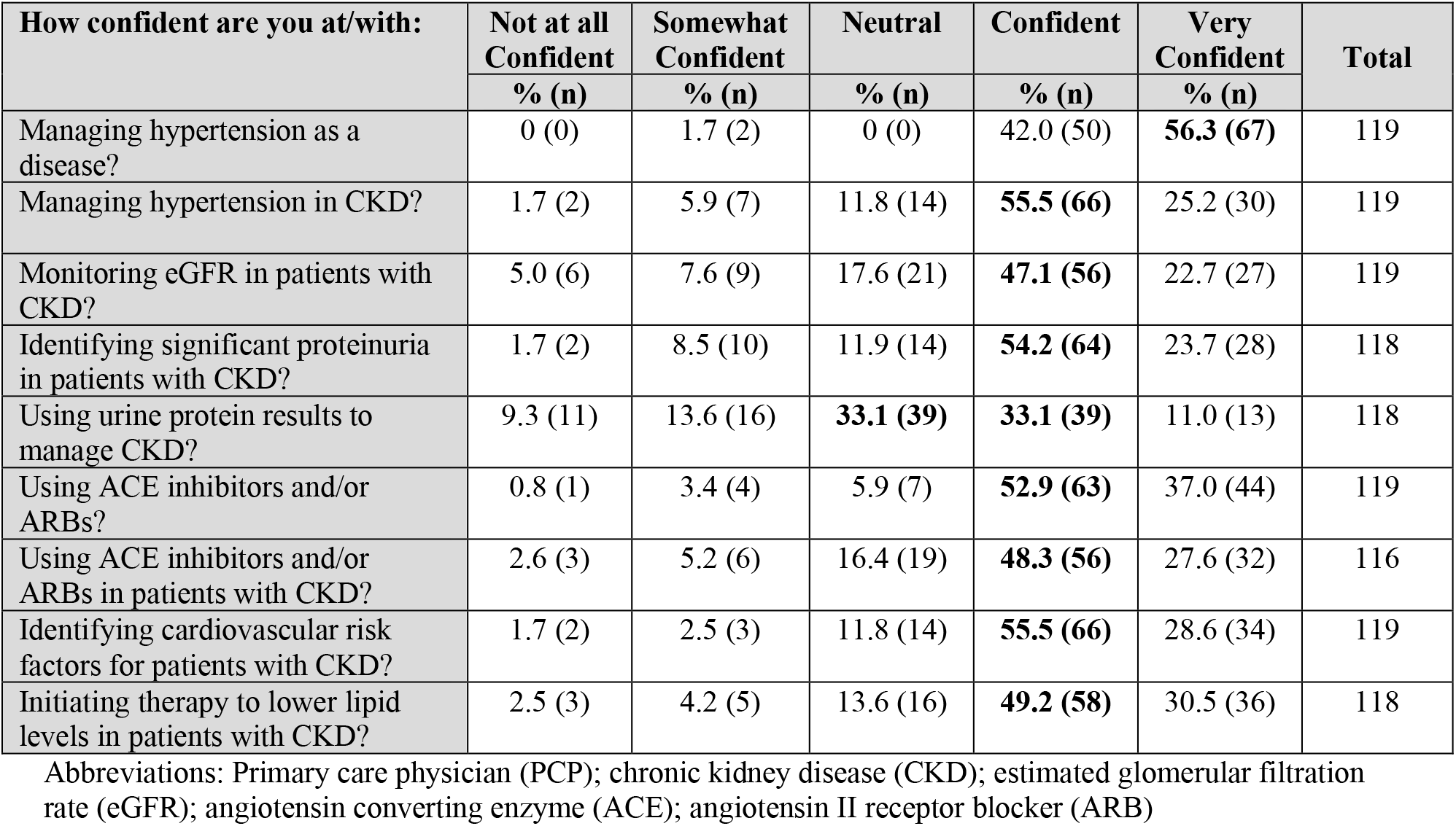
PCPs’ Confidence in Monitoring and Managing CKD.

| How confident are you at/with: | Not at all Confident | Somewhat Confident | Neutral | Confident | Very Confident | Total |
| --- | --- | --- | --- | --- | --- | --- |
|  | % (n) | % (n) | % (n) | % (n) | % (n) |  |
| Managing hypertension as a disease? | 0 (0) | 1.7 (2) | 0 (0) | 42.0 (50) | **56.3 (67)** | 119 |
| Managing hypertension in CKD? | 1.7 (2) | 5.9 (7) | 11.8 (14) | **55.5 (66)** | 25.2 (30) | 119 |
| Monitoring eGFR in patients with CKD? | 5.0 (6) | 7.6 (9) | 17.6 (21) | **47.1 (56)** | 22.7 (27) | 119 |
| Identifying significant proteinuria in patients with CKD? | 1.7 (2) | 8.5 (10) | 11.9 (14) | **54.2 (64)** | 23.7 (28) | 118 |
| Using urine protein results to manage CKD? | 9.3 (11) | 13.6 (16) | **33.1 (39)** | **33.1 (39)** | 11.0 (13) | 118 |
| Using ACE inhibitors and/or ARBs? | 0.8 (1) | 3.4 (4) | 5.9 (7) | **52.9 (63)** | 37.0 (44) | 119 |
| Using ACE inhibitors and/or ARBs in patients with CKD? | 2.6 (3) | 5.2 (6) | 16.4 (19) | **48.3 (56)** | 27.6 (32) | 116 |
| Identifying cardiovascular risk factors for patients with CKD? | 1.7 (2) | 2.5 (3) | 11.8 (14) | **55.5 (66)** | 28.6 (34) | 119 |
| Initiating therapy to lower lipid levels in patients with CKD? | 2.5 (3) | 4.2 (5) | 13.6 (16) | **49.2 (58)** | 30.5 (36) | 118 |

Abbreviations: Primary care physician (PCP); chronic kidney disease (CKD); estimated glomerular filtration rate (eGFR); angiotensin converting enzyme (ACE); angiotensin II receptor blocker (ARB)

Statistical differences of responses were assessed for comfort in diagnosing and managing various aspects of CKD within medical specialty area, medical training, practice setting and following CKD guidelines, see Table S3 in Supplemental Material. For medical specialty area, statistically significant differences were seen with all aspects except making the diagnosis of CKD in patients (p = 0.08). For medical training, no statistical differences were seen for bone disorders (p = 0.215), electrolyte disorders (p = 0.116) and metabolic acidosis (p = 0.793). Statistically significant differences were seen in following CKD guidelines for most aspects and when compared to medical specialty and training, it showed statistical significance with making a diagnosis of CKD (p = 0.013), managing bone disorders (p = 0.033), electrolyte disorders (p = 0.014) and metabolic acidosis (p = 0.021).

Statistical differences of responses were assessed for confidence in monitoring and managing CKD. Within medical training, statistically significant differences of responses were only seen for monitoring eGFR (p = 0.000), identifying cardiovascular risk factors (p = 0.001), and initiating lipid lowering therapy for patients with CKD (p = 0.001). Analysis of whether physicians followed CKD guidelines revealed statistically significant differences for all aspects of confidence in monitoring and managing CKD (p = 0.000 – 0.041), see Table 2 below.

**Table 2:**
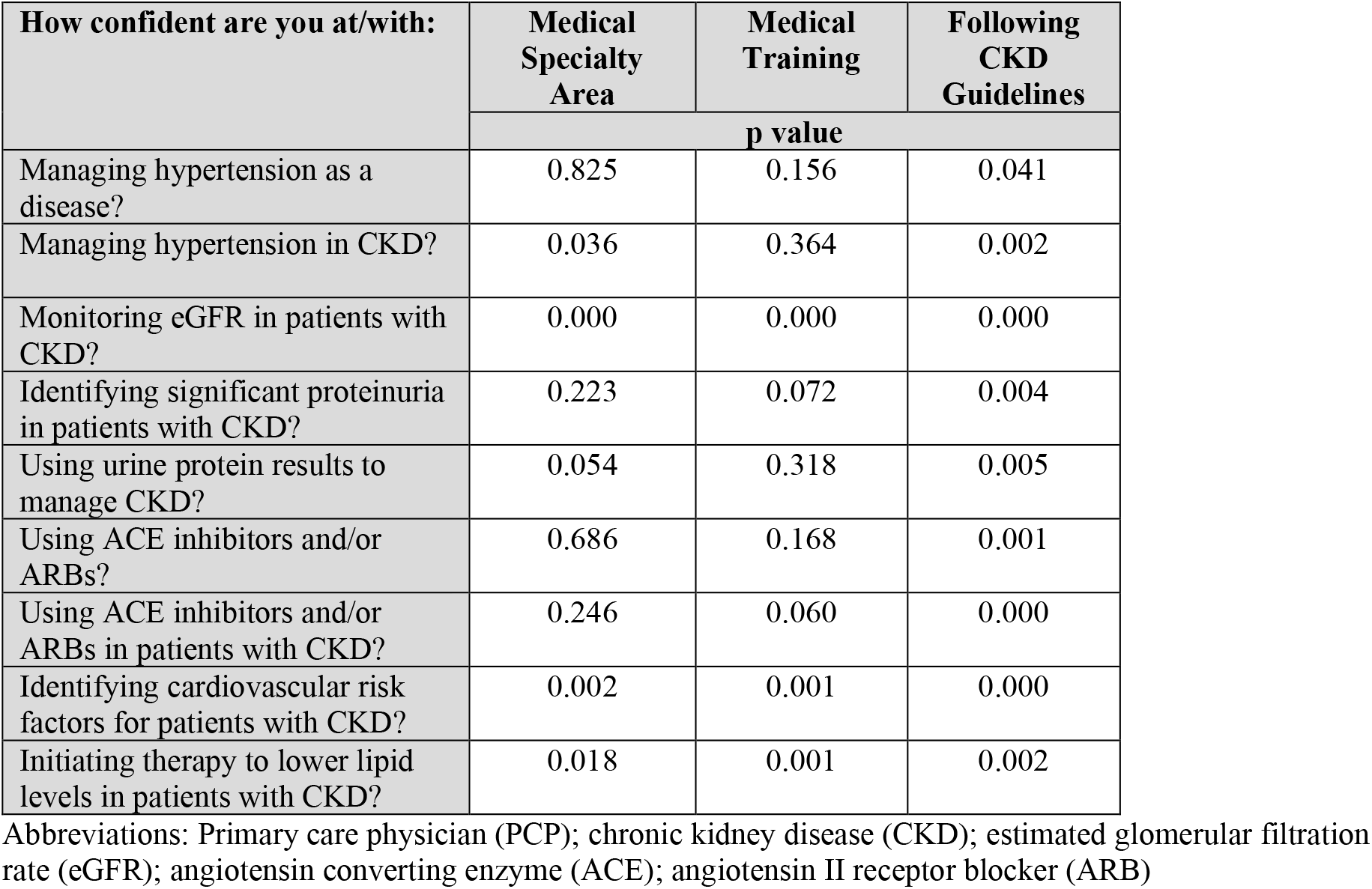
Statistical Significance of PCPs’ Confidence in Monitoring and Managing CKD.

| How confident are you at/with: | Medical Specialty Area | Medical Training | Following CKD Guidelines |
| --- | --- | --- | --- |
|  | p value |  |  |
| Managing hypertension as a disease? | 0.825 | 0.156 | 0.041 |
| Managing hypertension in CKD? | 0.036 | 0.364 | 0.002 |
| Monitoring eGFR in patients with CKD? | 0.000 | 0.000 | 0.000 |
| Identifying significant proteinuria in patients with CKD? | 0.223 | 0.072 | 0.004 |
| Using urine protein results to manage CKD? | 0.054 | 0.318 | 0.005 |
| Using ACE inhibitors and/or ARBs? | 0.686 | 0.168 | 0.001 |
| Using ACE inhibitors and/or ARBs in patients with CKD? | 0.246 | 0.060 | 0.000 |
| Identifying cardiovascular risk factors for patients with CKD? | 0.002 | 0.001 | 0.000 |
| Initiating therapy to lower lipid levels in patients with CKD? | 0.018 | 0.001 | 0.002 |

Abbreviations: Primary care physician (PCP); chronic kidney disease (CKD); estimated glomerular filtration rate (eGFR); angiotensin converting enzyme (ACE); angiotensin II receptor blocker (ARB)

A 5-point Likert scale was used to assess physicians’ agreement with 13 perceived barriers to the management of CKD, see Table S4 in Supplemental Material. Of the 4 patient-level barriers, physicians “agreed – strongly agreed”, with patient’s limited understanding (87.4%, n = 104), financial difficulties (84.0%, n = 100), non-adherence with medical management (89.9%, n = 107), and fears and/or beliefs (87.4%, n = 104) being barriers to CKD management. For the 4 provider-level barriers, physicians “agreed – strongly agreed” with PCPs limited recognition about CKD (49.2%, n = 58), lack of awareness of CKD guidelines (54.3%, n = 64), and risk factors being difficult to manage (49.6%, n = 59) as barriers, and “disagreed – strongly disagreed” with PCPs’ belief that they are unable to improve CKD (45.7%, n = 54). For the 5 systems-level barriers, physicians “agreed – strongly agreed” with limited time visit (65.3%, n = 77), lack of clinical information systems (60.5%, n = 72), insufficient support tools for patients (67.0%, n = 79), long wait times for specialty care services (79.9%, n = 95), and delays in completing investigations (73.9%, n = 88) as barriers to the management of CKD.

Participants were given an opportunity to state unique barriers to the management of CKD that they have encountered during their practice as a PCP. There were 26 unique barriers expressed, which were divided into patient-, provider-, and systems-level categories. Of the 8 patient-level barriers, the most common were non-adherence with nephrology appointments, lack of support (family and community) for patients with CKD, and uncontrolled comorbidities due to patient burnout. Of the 7 provider-level barriers, the most common were limited local CKD continuous medical education, lack of communication between PCP and nephrologist, and lack of continuity of care at community clinics. There were 11 unique systems-level barriers, with difficulty of the referral process to the public system, access to tests to diagnose CKD, and issues with the Princess Margaret Hospital Laboratory services being the most common barriers.

## Discussion

In this study PCPs’ evaluation, and management practices of patients with CKD were assessed, and barriers affecting overall care were determined. Over two-thirds of the PCP population for New Providence participated in this study with most physicians being from Family Medicine and General Practice medical specialty areas. The population of Internal Medicine physicians in primary care was much smaller than expected accounting for less than 10% of the sample frame; however, half of that Internal Medicine physician population participated in the study. Issues in the evaluation of patients with CKD were seen with the identification of at-risk groups, low utilization of urine diagnostic studies, and staging of CKD. For management practices, issues were seen with bone disorders, metabolic acidosis, and proteinuria in patients with CKD. Numerous perceived and unique patient-, provider-, and systems-level barriers were identified.

Evidence-based practice has become a standard of care among healthcare professionals and this study showed that overall, two-thirds of physicians expressed following 1 or more CKD guidelines; additionally, more than three-quarters of physicians in Family Medicine and Internal Medicine reported following CKD guidelines, compared to less than half of physicians in General Practice. This suggested that physicians who are in or have completed training programs tend to follow CKD guidelines compared to physicians who are not in training programs. In Sperati et al., physicians mostly reported seeing > 10 CKD patients per week^5^, which differs from our study where physicians reported seeing ≤ 10.

Some physicians may have a small number of CKD patients within their practice, and lack of exposure with CKD patients can impact the evaluation and management of current and future patients, thus leading to progression of disease.

In our study, the least commonly selected at-risk groups for CKD were elderly, smoking and obesity with similar findings seen in Delatorre et al^6^. According to the World Health Organization, “by 2030, 1 in 6 people will be aged 60 years and over”, and in The Bahamas STEPS Survey (2019), 43.7% of the study participants were obese and the proportion of current smokers and those with secondhand exposure had increased from the previous STEPS survey.^7,8^ With the world’s population aging, and obesity and smoking still being risk factors in our population, PCPs in our study are potentially neglecting to screen patients because they fall into a category that they do not consider to be at risk of CKD. The diagnostic tests used to identify patients with CKD varied amongst the physicians in this study; however, urine testing was selected less frequently. Patients can have significant proteinuria while having normal serum creatinine and eGFR, and as such points to a group of CKD patients that may be unidentified in our population. In Tu et al., review of >140,000 electronic medical records revealed uACR testing was the most poorly used indicator of CKD with less than half of patients at risk for CKD having one.^9^ Lack of using urine studies suggested physicians may be unaware of this subset of CKD patients and indicates why staging of CKD was commonly reported as using eGFR alone versus eGFR and uACR in our study. It may also be due to lack of knowledge regarding interpretation of results, as well as patient financial constraints in getting the urine tests done.

Physicians were overall comfortable diagnosing and managing CKD and its complications except for bone disorders and metabolic acidosis where physicians were mostly neutral and had lower levels of agreement (disagree – strongly disagree). Physicians also cited lack of available tools to help them and their patients with these CKD complications, which may reflect a lack of exposure and education, suggesting a need to provide more resources on these specific complications. Physicians in FM and IM specialty areas, physicians with Doctor of Medicine (DM) in FM/IM, and physicians following CKD guidelines had higher levels of agreement (agreed – strongly agreed) with comfort in diagnosing and managing CKD and its complications compared to GPs, physicians with non-DM specialty degrees, and physicians not following CKD guidelines. This suggests that training and education has a positive impact on physicians’ comfort with CKD care.

Physicians were overall confident with monitoring and managing CKD, however they were equally neutral and confident with using protein results to manage CKD, which further suggests that lack of knowledge and usage with urine protein studies.

Many barriers to the management of CKD in our population were revealed. Physicians strongly agreed with 3 perceived barriers; i. Patients unable to afford recommended CKD care, ii. Patients’ non-adherence with medical management, and iii. Long wait-times for specialty care services. The COVID-19 pandemic led to persons being temporarily or permanently unemployed which caused new or worsened financial constraints, making it less likely for patients to afford medications and adhere to regimes. It also created further delays in outpatient follow-ups for specialty care clinics due to the temporary discontinuation of face-to-face visits. Of the unique patient-level barriers, non-adherence with nephrology appointments was frequently stated. Patients in some instances would decline visits with the nephrologists, despite being informed about their CKD status by their PCP which could be due to their limited understanding or from fears and/or beliefs about CKD. Of the unique provider-level barriers, limited local CKD continuous medical education (CME), lack of communication between PCP and nephrologist, and lack of continuity of care at community clinics were frequently stated. Physicians agreed from the perceived barriers that PCPs had limited recognition/knowledge about CKD and lacked awareness of CKD guidelines. Therefore, having access to local CMEs provided by our nephrologists can begin to bridge gaps in CKD care provided by our PCPs. Lack of continuity of care at community clinics occurs from the unpredictable shift schedules and unexpected location changes due to staff shortages within the public sector. This can lead to deficiencies in effective care of chronic non-communicable diseases such as CKD. Of the unique systems-level barriers, difficulty of the referral process to the public system was frequently reported. Within the public sector, referral letters are only accepted on public forms, therefore if a patient is seen privately and needs referral to a public specialty service, the private form would have to be re-written on a public form and/or they would have to be seen by a public sector physician to facilitate the referral. This leads to increased patient volumes at the community clinics, and increased frustration for both the patients and physicians, which ultimately leads to delays in the evaluation and management of CKD.

Study limitations included the lack of an official list of primary care physicians in New Providence which made the process of exclusion more tedious than expected. Based on the results of this study, recommendations include the development of a National CKD guideline, local CKD continuous medical education, implementation of an electronic referral and appointment platform to connect the public and private sectors, and CKD public health education for patients. In conclusion, this study highlighted deficits in the evaluation and management of CKD with respect to proteinuria and specific complications such as bone disorders and metabolic acidosis. It also identified numerous patient-, provider-, and systems-level barriers to the overall care of patients with CKD. Future quantitative and qualitative studies are needed to further explore CKD at the primary care level and incorporate views of our nephrologists.

## Data Availability

All data produced in the present study are available upon reasonable request to the authors

## Declarations

### Data availability

Research records will be kept for at least 5 years from the time data collection was completed (February 2023). Readers may access the data on which the analysis and conclusion of the study were based upon justified request from the corresponding author.

### Authors’ contributions

*Wilnaye Bain:* Conceptualization, Data curation, Formal analysis, Investigation, Methodology, Writing – original draft preparation, Writing – review & editing.

*Sabriquet Pinder-Butler:* Supervision, Writing – review & editing.

*Terrance Fountain:* Formal analysis, Methodology, Software, Validation.

*Ilsa Grant:* Validation, Supervision, Writing – review & editing.

## Acknowledgements

We would like to extend our sincere thanks to the following people that were influential with the generation of this research.

Dr. Morton A. Frankson, Epidemiologist & Lecturer, The University of the West Indies School of Clinical Medicine & Research (UWISCMR), provided guidance and support during the proposal phase of this research.

Our research assistants, Dr. Merrilyn Wallace-Bain, and Dr. Briquel Sherman-Knowles who were willing to provide support during the data collection process.

Ms. Suja Philip, Librarian, UWISCMR, who assisted in acquiring articles during the literature review process.

We would also like to thank the physicians who participated in this study.

