## Supplemental Material for "Primary Care Physicians’ Practices and Barriers in Evaluating and Managing Chronic Kidney Disease in New Providence, The Bahamas"

### Supplemental Data

Table S1: Demographic Profile of Study Participants

| CHARACTERISTIC | % (n) | CHARACTERISTIC | % (n) |
|---|---|---|---|
| **Age** | | **Practice Setting** | |
| Mean: 45.1 ± 10.1 years | - | Public service only | 29.4 (35) |
| **Gender** | | Private Practice only | 36.1 (43) |
| Male | 36.1 (43) | Public and private practice | 34.5 (41) |
| Female | 63.9 (76) | **Public Service Department** | |
| **Ethnicity** | | Department of Family Medicine | 55.1 (38) |
| Black/Afro-Caribbean | 91.5 (108) | Department of Public Health | 34.8 (24) |
| White | 1.7 (2) | Department of Internal Medicine | 10.1 (7) |
| Asian | 4.2 (5) | **Years in Practice** | |
| Hispanic/Latino | 1.7 (2) | Mean: 15.8 ± 10.3 years | - |
| Other | 0.8 (1) | **Number of patients/week** | |
| **Medical Specialty Area** | | 50 or less | 28.6 (34) |
| General Practice | 41.2 (49) | 51-100 | 51.3 (61) |
| Family Medicine | 52.1 (62) | 101-150 | 16.0 (19) |
| Internal Medicine | 6.7 (8) | 151-200 | 3.4 (4) |
| **Medical Training** | | >200 | 0.8 (1) |
| MBBS | 31.4 (37) | **Number of CKD patients/week** | |
| MD | 12.7 (15) | 10 or less | 73.7 (87) |
| Diploma in Family Medicine | 12.7 (15) | 11-20 | 25.4 (30) |
| MSc in Family Medicine | 4.2 (5) | 21-30 | 0.8 (1) |
| DM in Family Medicine | 28.8 (34) | **Follow CKD guidelines** | |
| DM in Internal Medicine | 5.9 (7) | No | 32.1 (35) |
| Other | 4.2 (5) | Yes | 67.9 (74) |

Table S2.1: PCPs' Agreement with Comfort in Diagnosing and Managing CKD

| I feel comfortable: | Strongly Disagree | Disagree | Neutral | Agree | Strongly Agree | Total |
|---|---|---|---|---|---|---|
| | % (n) | % (n) | % (n) | % (n) | % (n) | |
| Making the diagnosis of CKD in my patients | 0.8 (1) | 2.5 (3) | 7.6 (9) | 43.7 (52) | **45.4 (54)** | 119 |
| Educating my patients about CKD | 0 (0) | 3.4 (4) | 8.4 (10) | **50.4 (60)** | 37.8 (45) | 119 |
| Managing my patients with CKD | 1.7 (2) | 7.6 (9) | 23.5 (28) | **48.7 (58)** | 18.5 (22) | 119 |
| Managing medication dosing in my patients with CKD | 0.9 (1) | 10.3 (12) | 17.1 (20) | **48.7 (57)** | 23.1 (27) | 117 |
| Avoiding nephrotoxic medications (e.g., NSAIDs) in my patients with CKD | 0.8 (1) | 0.8 (1) | 9.3 (11) | **46.6 (55)** | 42.4 (50) | 118 |
| Managing anemia of CKD in my patients | 1.7 (2) | 7.6 (9) | 29.7 (35) | **47.5 (56)** | 13.6 (16) | 118 |
| Managing bone disorders of CKD in my patients | 6.8 (8) | 23.7 (28) | **43.2 (51)** | 22.0 (26) | 4.2 (5) | 118 |
| Managing electrolyte disorders (e.g., hyperkalemia) in my patients with CKD | 4.2 (5) | 10.1 (12) | 30.3 (36) | **40.3 (48)** | 15.1 (18) | 119 |
| Managing metabolic acidosis in my patients with CKD | 5.1 (6) | 23.7 (28) | **34.7 (41)** | 24.6 (29) | 11.9 (14) | 118 |

Table S2.2: PCPs' Agreement with Availability of Tools to Diagnose and Manage CKD

| I have available tools (e.g., checklists, printed/web-based/smartphone-based resources) which help me to: | Strongly Disagree | Disagree | Neutral | Agree | Strongly Agree | Total |
|---|---|---|---|---|---|---|
| | % (n) | % (n) | % (n) | % (n) | % (n) | |
| Diagnose CKD | 3.4 (4) | 2.5 (3) | 2.5 (3) | **46.2 (55)** | 45.4 (54) | 119 |
| Manage CKD | 4.2 (5) | 5.1 (6) | 11.0 (13) | **48.3 (57)** | 31.4 (37) | 118 |
| Manage medication dosing | 3.4 (4) | 3.4 (4) | 7.6 (9) | **49.6 (59)** | 36.1 (43) | 119 |
| Avoid prescribing nephrotoxic medications | 1.7 (2) | 2.5 (3) | 6.7 (8) | **55.5 (66)** | 33.6 (40) | 119 |
| Manage hypertension in my patients with CKD | 0.8 (1) | 0.8 (1) | 6.7 (8) | **55.5 (66)** | 36.1 (43) | 119 |
| Manage anemia of CKD | 0.8 (1) | 9.3 (11) | 24.6 (29) | **46.6 (55)** | 18.6 (22) | 118 |
| Manage bone disorders of CKD | 4.2 (5) | 21.8 (26) | **35.3 (42)** | 29.4 (35) | 9.2 (11) | 119 |
| Manage hyperkalemia in CKD | 2.5 (3) | 11.8 (14) | 24.4 (29) | **47.1 (56)** | 14.3 (17) | 119 |
| Manage metabolic acidosis in CKD | 4.2 (5) | 19.3 (23) | **31.9 (38)** | **31.9 (38)** | 12.6 (15) | 119 |

Table S2.3: PCPs' Agreement with Availability of Tools to Help CKD Patients

| **I have educational tools and resources (e.g., printed and web-based materials/programs, classes, or health educators) available to help my patients understand:** | **Strongly Disagree** | **Disagree** | **Neutral** | **Agree** | **Strongly Agree** | **Total** |
|---|---|---|---|---|---|---|
| | **% (n)** | **% (n)** | **% (n)** | **% (n)** | **% (n)** | |
| Their CKD diagnosis | 4.2 (5) | 19.3 (23) | 10.1 (12) | **45.4 (54)** | 21.0 (25) | 119 |
| The potential medication-related risks associated with CKD (e.g., nephrotoxins) | 5.0 (6) | 19.3 (23) | 13.4 (16) | **43.7 (52)** | 18.5 (22) | 119 |
| Anemia of CKD | 4.2 (5) | 22.0 (26) | 19.5 (23) | **38.1 (45)** | 16.1 (19) | 118 |
| Hypertension in CKD | 3.4 (4) | 17.6 (21) | 16.0 (19) | **41.2 (49)** | 21.8 (26) | 119 |
| Bone disorders in CKD patients | 5.1 (6) | **32.2 (38)** | 28.8 (34) | 23.7 (28) | 10.2 (12) | 118 |
| Hyperkalemia in CKD | 5.9 (7) | 23.5 (28) | 25.2 (30) | **33.6 (40)** | 11.8 (14) | 119 |
| Metabolic acidosis in CKD | 5.0 (6) | **32.8 (39)** | 24.4 (29) | 27.7 (33) | 10.1 (12) | 119 |

Table S3: Statistical Significance of PCPs' Comfort in Diagnosing and Managing CKD

| **I feel comfortable:** | **Medical Specialty Area** | **Medical Training** | **Practice Setting** | **Following CKD Guidelines** |
|---|---|---|---|---|
| | **p value** | | | |
| Making the diagnosis of CKD in my patients | 0.080 | 0.006 | 0.344 | 0.013 |
| Educating my patients about CKD | 0.001 | 0.007 | 0.283 | 0.005 |
| Managing my patients with CKD | 0.000 | 0.001 | 0.006 | 0.000 |
| Managing medication dosing in my patients with CKD | 0.000 | 0.006 | 0.010 | 0.015 |
| Avoiding nephrotoxic medications (e.g., NSAIDs) in my patients with CKD | 0.035 | 0.034 | 0.119 | 0.103 |
| Managing anemia of CKD in my patients | 0.009 | 0.035 | 0.197 | 0.074 |
| Managing bone disorders of CKD in my patients | 0.015 | 0.215 | 0.302 | 0.033 |
| Managing electrolyte disorders (e.g., hyperkalemia) in my patients with CKD | 0.000 | 0.116 | 0.004 | 0.014 |
| Managing metabolic acidosis in my patients with CKD | 0.000 | 0.793 | 0.012 | 0.021 |

Table S4: PCPs' Perceived Barriers to the Overall Care of CKD Patients

| **PERCEIVED BARRIERS:** | **Strongly Disagree** | **Disagree** | **Neutral** | **Agree** | **Strongly Agree** | **Total** |
|---|---|---|---|---|---|---|
| | **% (n)** | **% (n)** | **% (n)** | **% (n)** | **% (n)** | |
| Patients' limited understanding about CKD and its' implications | 0 (0) | 5.0 (6) | 7.6 (9) | **47.9 (57)** | 39.5 (47) | 119 |
| Patients unable to afford recommended CKD care (e.g., tests, medications) | 0.8 (1) | 3.4 (4) | 11.8 (14) | 37.8 (45) | **46.2 (55)** | 119 |
| Primary care physicians' limited recognition or knowledge about CKD | 4.2 (5) | 16.1 (19) | 30.5 (36) | **39.0 (46)** | 10.2 (12) | 118 |
| Primary care physicians' lack of awareness of CKD guidelines or useful algorithms for CKD care | 4.2 (5) | 13.6 (16) | 28.0 (33) | **40.7 (48)** | 13.6 (16) | 118 |
| CKD risk factors (e.g., HTN, DM, Obesity) are difficult to manage | 10.1 (12) | 28.6 (34) | 11.8 (14) | **41.2 (49)** | 8.4 (10) | 119 |
| Primary care physicians' belief that they are unable to improve CKD | 7.6 (9) | **38.1 (45)** | 28.8 (34) | 19.5 (23) | 5.9 (7) | 118 |
| Limited visit time to care for complex patients | 3.7 (4) | 11.9 (14) | 19.5 (23) | **49.2 (58)** | 16.1 (19) | 118 |
| Lack of comprehensive clinical information systems – EMR (e.g., patient registries) | 2.5 (3) | 11.8 (14) | 25.2 (30) | **37.8 (45)** | 22.7 (27) | 119 |
| Insufficient clinical support tools and resources to support patient self-management | 0.8 (1) | 15.3 (18) | 16.9 (20) | **42.4 (50)** | 24.6 (29) | 118 |
| Patient's non-adherence with medical management (e.g., medications to control comorbidities, lifestyle changes) | 0 (0) | 0 (0) | 10.1 (12) | 42.0 (50) | **47.9 (57)** | 119 |
| Patient's fears and/or beliefs about management for end-stage renal disease (e.g., dialysis, kidney transplantation) | 0 (0) | 3.4 (4) | 9.2 (11) | **46.2 (55)** | 41.2 (49) | 119 |
| Long wait times for specialty care services (e.g., nephrology clinic, vascular clinic) | 0.8 (1) | 9.2 (11) | 10.1 (12) | 37.0 (44) | **42.9 (51)** | 119 |
| Delays in completing investigations to diagnose CKD (e.g., wait time for ultrasound scan of kidneys) | 1.7 (2) | 11.8 (14) | 12.6 (15) | **42.0 (50)** | 31.9 (38) | 119 |
